# Clinical value of the fibrosis-4 index in predicting mortality in patients with right ventricular pacing

**DOI:** 10.1101/2023.10.30.23297769

**Authors:** Naoya Inoue, Shuji Morikawa, Takashi Ogane, Takehiro Hiramatsu, Toyoaki Murohara

## Abstract

**Background:** The fibrosis-4 (FIB-4) index has attracted attention as a predictive factor for cardiovascular events and mortality in patients with heart disease. However, its clinical value in patients with implanted pacemakers remains unclear.

**Methods:** This study included patients who underwent pacemaker implantation. The FIB-4 index was calculated based on blood tests performed during the procedure. The primary outcome was all-cause mortality, and secondary outcomes included cardiovascular and non-cardiovascular death. The FIB-4 index was stratified into tertiles. Between-group comparisons were performed using log-rank tests and multivariate analysis using Cox proportional hazards. The predictive accuracy and cut-off value of the FIB-4 index were calculated from the receiver operating characteristic curve for all-cause mortality.

**Results:** This study included 201 participants, of whom 38 (incidence rate: 5.8/100 person-years) experienced death events during the observation period (median: 1097 days). All-cause and non-cardiovascular death differed significantly between groups stratified by the FIB-4 index tertiles (log-rank test: *P*<0.001 and *P*<0.001, respectively). Using Cox proportional hazards analysis, the unadjusted hazard ratio was 4.75 (95% confidence interval [CI]: 2.05–11.0, *P*<0.001) for Tertile 3 compared to Tertile 1. After adjustment for confounding factors, including age, sex, the presence or absence of left bundle branch block at baseline, QRS duration during pacing, and pacing rate at the last check, the hazard ratio was 3.61 (95% CI: 1.37–9.48, *P*=0.009). The cut-off of the FIB-4 index was 3.75 (area under the curve: 0.72, 95% CI: 0.62–0.82).

**Conclusions:** The FIB-4 index was a potential predictive factor for all-cause mortality in patients with implanted pacemakers.

## Introduction

In the past, right ventricular pacing (RVP) was used for bradycardia arrhythmias; however, this was associated with problems like decreased left ventricular systolic function [1], increased hospitalization for heart failure [2], and the onset of atrial fibrillation [3] due to electrical and mechanical dyssynchrony. To address these issues, pacing techniques involving the His bundle or left bundle area have been recently introduced [4,5]. Many studies have compared the outcomes of these novel pacing techniques with those of conventional RVP [6], and investigated the factors contributing to the problems associated with RVP, with pacing rate and QRS duration reported as problematic factors [7,8]. However, to the best of our knowledge, studies on the relationship between RVP and mortality are limited. The fibrosis-4 (FIB-4) index, a non-invasive marker used to assess the degree of liver fibrosis, has been reported as a predictive factor for right ventricular function and major adverse cardiovascular events in patients with heart diseases, such as heart failure and tricuspid regurgitation [9,10]. In a study targeting heart failure in patients with preserved left ventricular systolic function, those with a FIB-4 index ≥ 3.11 had a 2.202-fold (95% confidence interval: 1.110–4.368) higher risk of major adverse cardiovascular events (composite of cardiovascular death, heart failure-related rehospitalization, non-fatal myocardial infarction, and non-fatal stroke) [10]. The FIB-4 index is related to cardiovascular events because prolonged hepatic congestion due to reduced blood flow and sodium diuresis leads to fluid retention and arterial sclerosis, which impair left ventricular diastolic function [10,11]. However, no study to date has investigated the prognosis of patients with right ventricular pacemakers in relation to the FIB-4 index. Therefore, this study aimed to investigate whether the FIB-4 index is associated with mortality in patients with right ventricular pacemakers. Patients requiring pacemaker implantation often have circulatory insufficiency and congestion at the time of bradycardia arrhythmia onset, and many have elevated B-type natriuretic peptide (BNP) levels. If the FIB-4 index can be confirmed to independently predict prognosis, regardless of the pacing factors, it may have high clinical value.

## Materials and Methods

### Patients

This retrospective, single-center, observational study included patients who had undergone right ventricular pacemaker implantation for sinus node dysfunction or atrioventricular block between April 2015 and December 2021. The following patients were excluded: 1) those who underwent stimulation conduction system pacemaker implantation, such as His bundle pacing or left bundle area pacing; 2) those with a ventricular pacing rate < 1% at the final pacemaker check; 3) those who did not undergo follow-up at our hospital after right ventricular pacemaker implantation; 4) those for whom right ventricular electrocardiograms were unobtainable (including only fusion waveforms); and 5) those who underwent VVI-pacemaker implantation for bradycardia-induced atrial fibrillation. Ultimately, 201 patients were included in the study.

The study was conducted in accordance with the standards of the Declaration of Helsinki and current ethical guidelines and was approved by the Ethics Committee on Medical Research of Chutoen General Medical Center (reference no: 1241230822). As this study was not a clinical trial, and the data were retrospectively and anonymously collected and analyzed, the requirement for patients’ written informed consent was waived.

### Data collection

The data were accessed for research purposes on August 23, 2023. The authors had access to information that could identify individual participants during and after data collection.

The FIB-4 index was calculated based on the patient’s age and blood test results at the time of pacemaker implantation using the following formula: FIB-4 index = age × aspartate aminotransferase [AST] level / (platelets × square root [alanine transaminase level]). The study participants were divided into three groups based on their FIB-4 index values: Tertile 1 (FIB-4 ≤ 2.17; n = 67), Tertile 2 (2.18 ≤ FIB-4 ≤ 3.28; n = 67), and Tertile 3 (FIB-4 ≥ 3.29; n = 67). Patient characteristics were compared between the groups.

### Primary and secondary outcomes

The primary outcome of this study was all-cause mortality, which included deaths due to cardiovascular disease, cancer, infections, and other causes. Secondary outcomes included cardiovascular death, which comprised deaths resulting from fatal arrhythmias, fatal myocardial infarction, heart failure, and stroke, as well as non-cardiovascular death, which involved deaths caused by non-cardiovascular factors like cancer, infections, and renal failure.

### Electrocardiogram analysis

The 12-lead electrocardiograms (ECGs) obtained preoperatively and during pacemaker checks were analyzed. ECGs were recorded at a paper speed of 25 mm/s and an amplitude of 10 mm/mV. Two cardiologists independently evaluated the following parameters from each ECG: 1) the heart rate; 2) the QRS duration (both during bradycardia, arrhythmias, and pacing); 3) the presence or absence of conduction abnormalities.

### Right ventricular pacemaker implantation and pacemaker check

Every patient underwent implantation of a dual-chamber pacemaker in the right ventricle (RV). The specific site for RV lead implantation was the RV septum, aimed for under fluoroscopic guidance. Nonetheless, the ultimate determination regarding lead placement was at the discretion of the principal doctor, considering optimal electrical parameters and lead stability.

Pacemaker assessments were conducted prior, and subsequent, to implantation. Post-discharge, patients received regular outpatient follow-ups at intervals of 6 to 9 months. During these visits, appropriate ECG and pacemaker parameters were duly documented.

### Statistical analysis

Regarding the statistical analysis, categorical variables were analyzed using the chi-square or Fisher’s exact tests, and presented as numbers and percentages. Continuous variables were analyzed using non-parametric tests (Kruskal–Wallis test), and, if necessary, the Mann–Whitney U test was performed with the significance level adjusted using the Bonferroni method for multiple comparisons. The occurrence of mortality events during the post-implantation follow-up period was computed, and the curves depicting the incidence of these events were compared between groups utilizing log-rank assessment.

Univariate analysis using Cox proportional hazards was conducted for both primary and secondary outcomes. Multivariate analysis, adjusted for age and sex, was performed to determine the primary outcomes. Furthermore, multivariate analysis using the forced entry method was performed for other potential factors influencing clinical outcomes, such as the pacing rate during RVP, QRS duration, estimated glomerular filtration rate (eGFR), BNP level, and tricuspid regurgitation pressure gradient (TR-PG).

Statistical significance was set at *P* < 0.05. All statistical analyses were performed using R with EZR software (Saitama Medical Center, Jichi Medical University, Saitama, Japan) [12]. Regarding missing data, only the samples with complete data for individual statistical analyses were included.

## Results

Among patients who underwent pacemaker implantation at our hospital between April 2015 and December 2021, 201 were included in the final analysis. In the comparison of the FIB-4 index tertiles, the median FIB-4 index values were as follows: Tertile 1, 1.80; Tertile 2, 2.66; and Tertile 3, 4.19 (*P* < 0.001). Significant differences were observed in various elements of the FIB-4 index, including age, AST, platelet count, eGFR, hemoglobin, low-density lipoprotein cholesterol, BNP, and dyslipidemia (Table 1).

**Table 1.**
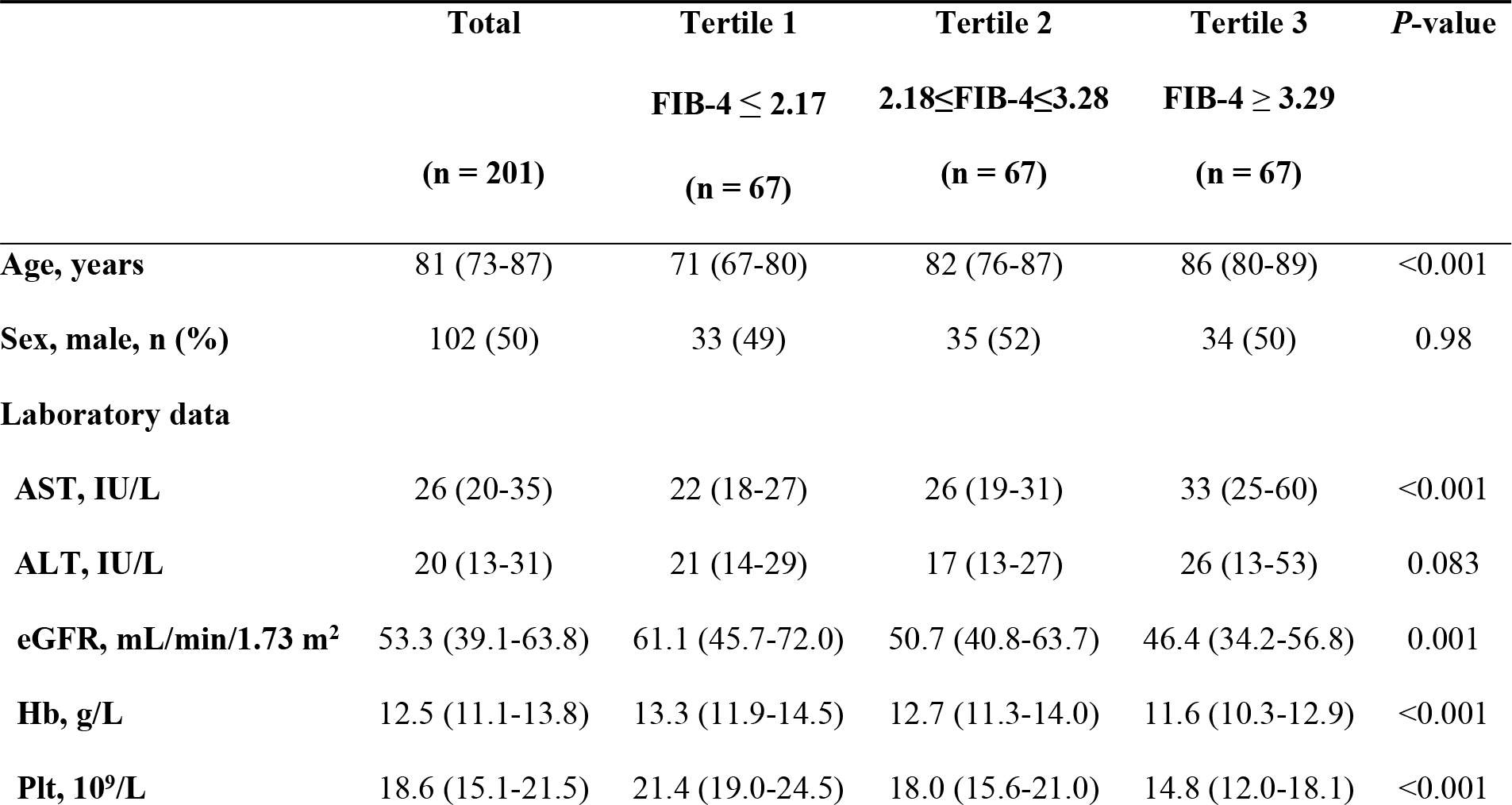
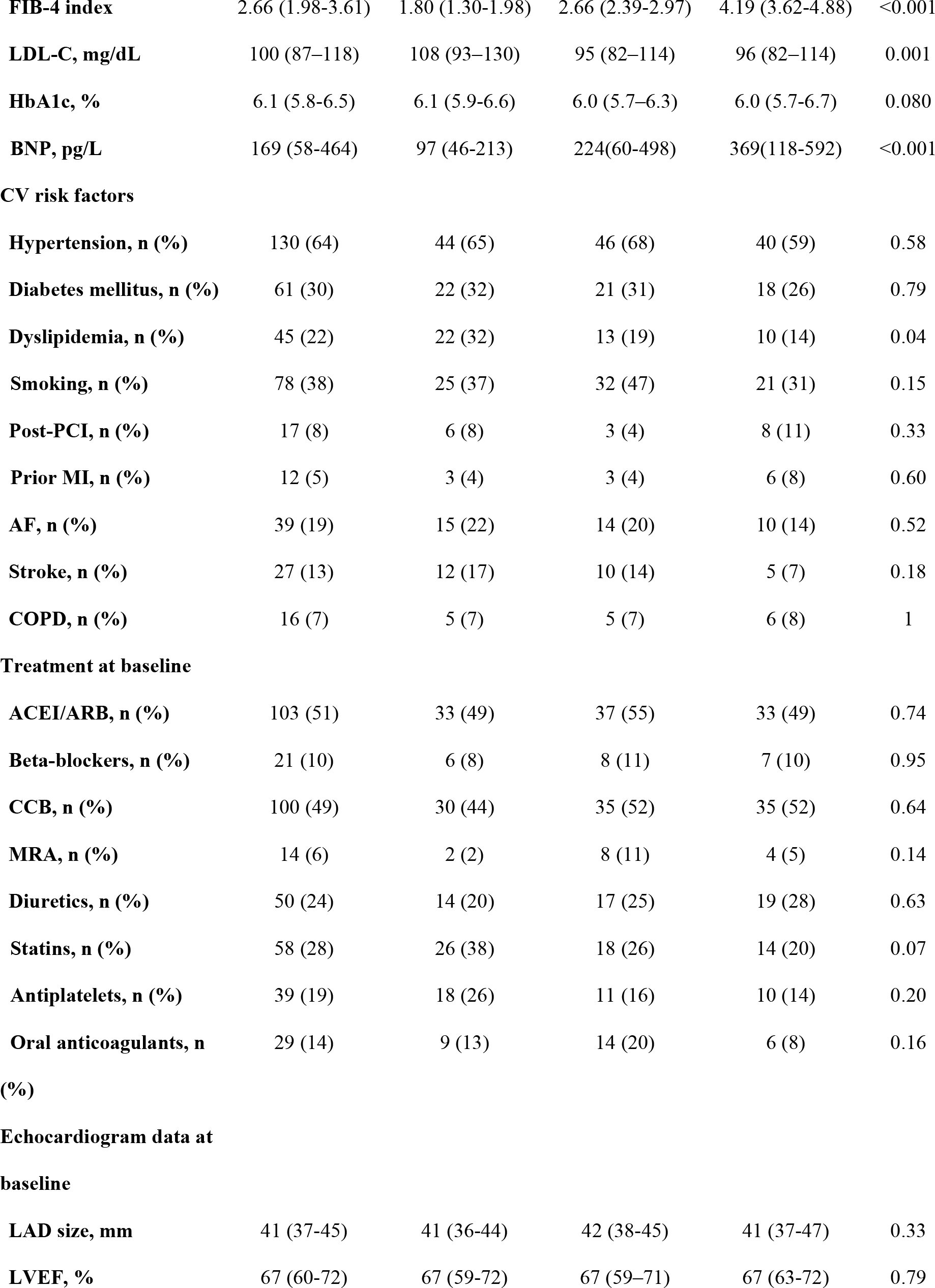
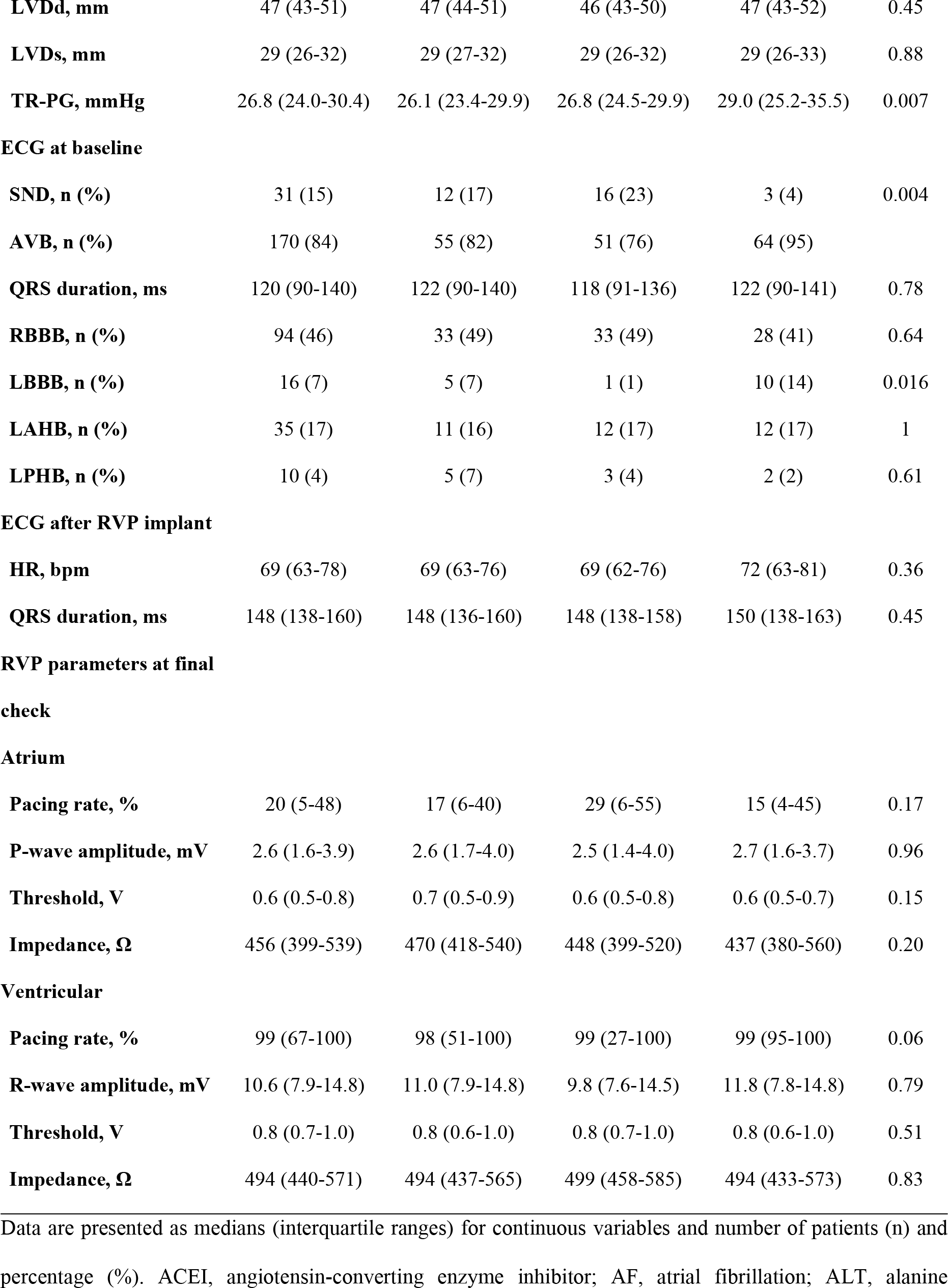
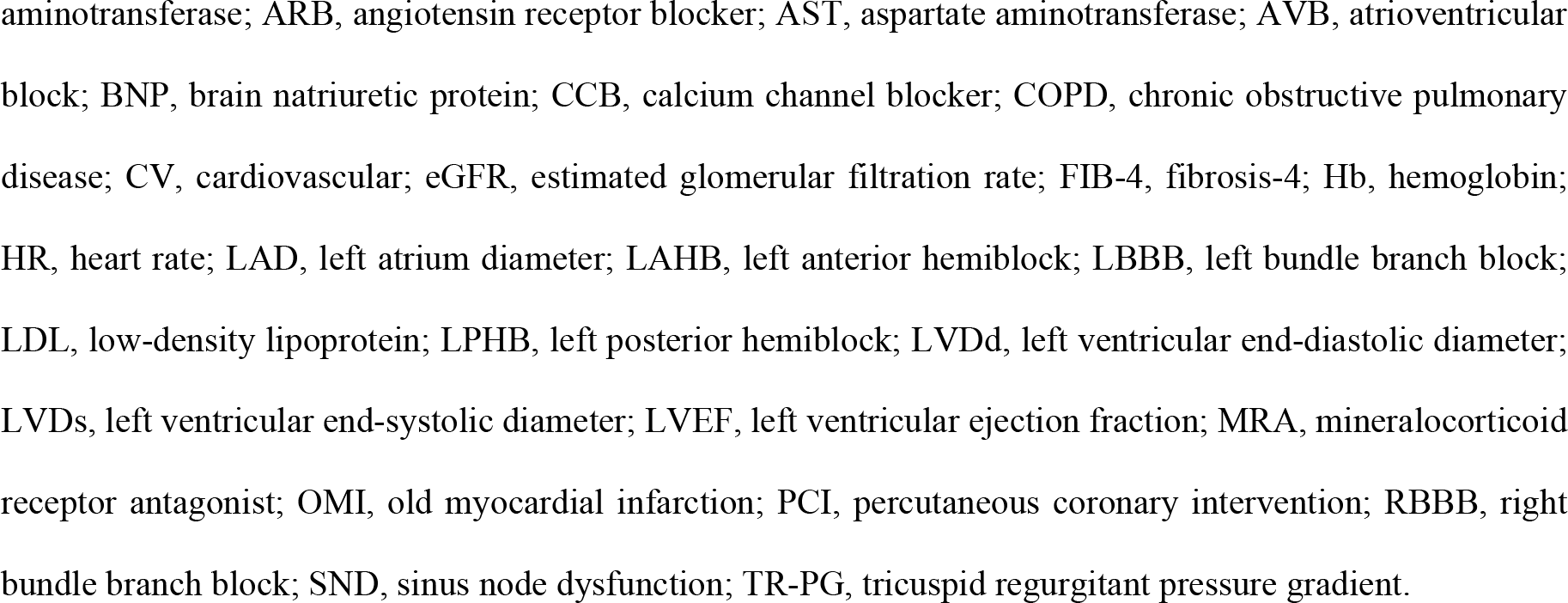
Patient characteristics according to the FIB-4 index tertiles.

No significant differences were observed in terms of baseline treatment, cardiac ultrasound examination results, or pacemaker parameters at the last follow-up. However, there was a significant difference in the proportion of left bundle branch block (LBBB) on the baseline ECG (Table 1).

Among the study patients (median age: 81 years), 38 died during the observation period (median: 1097 days), resulting in an incidence rate of 5.8/100 person-years. The incidence rates for each major outcome, stratified into three groups based on the FIB-4 index, were as follows: Tertile 1, 2.9/100 person-years; Tertile 2, 2.3/100 person-years; and Tertile 3, 13.3/100 person-years (log-rank test, *P* < 0.001; Fig 1A, Table 2).

**Table 2.**
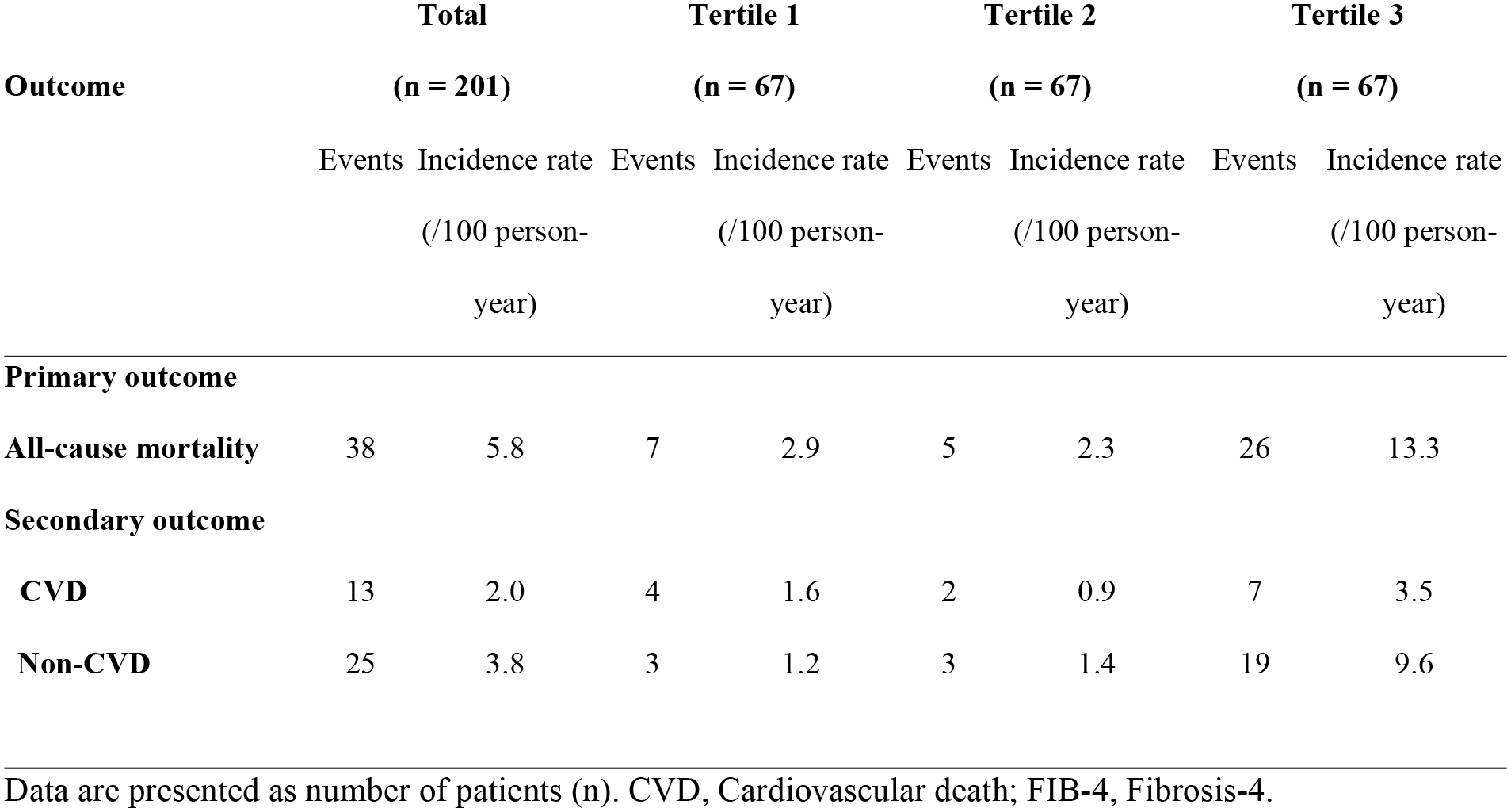
Primary and secondary outcomes.

**Fig 1.**
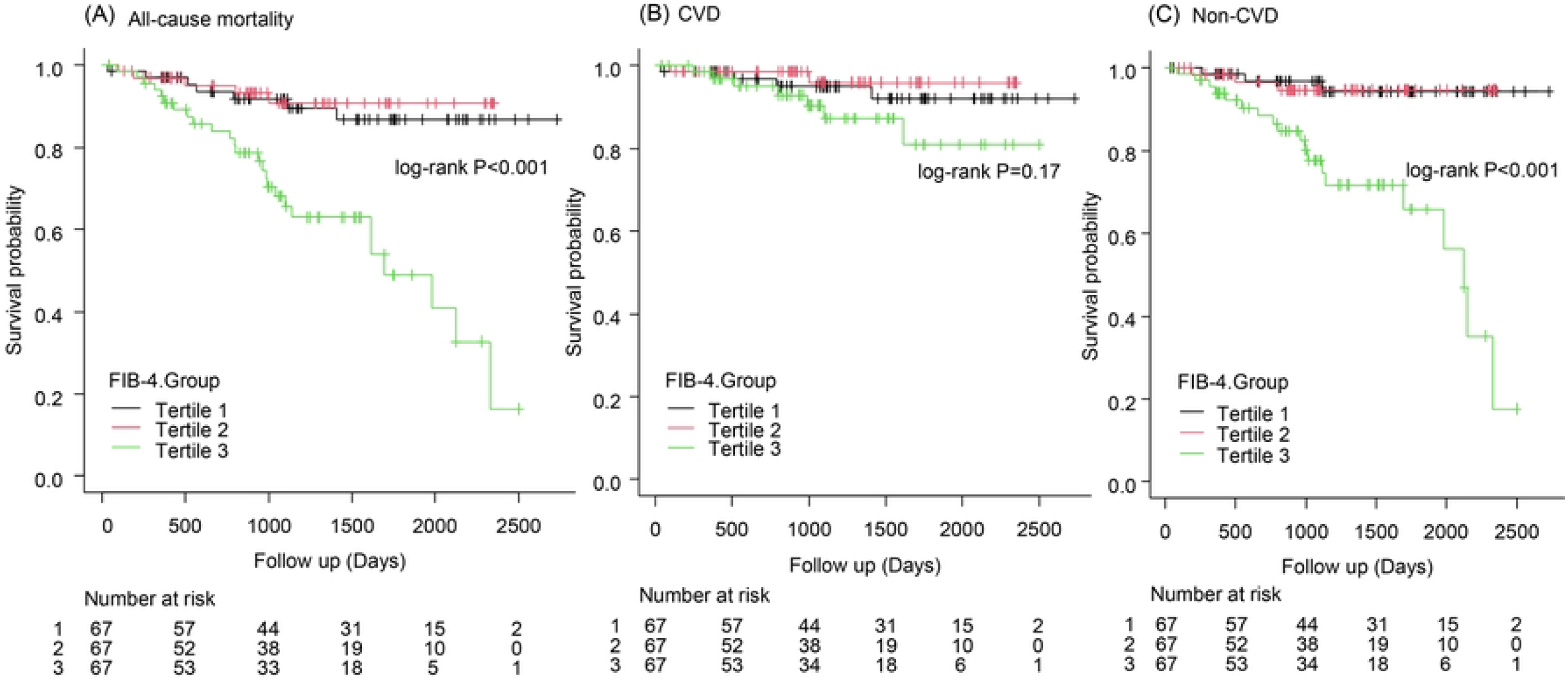
All-cause mortality and secondary outcomes stratified by FIB-4 index tertiles. (A) Kaplan–Meier curve of all-cause mortality. (B) Cardiovascular death. (C) Non-cardiovascular death. CVD, cardiovascular death; FIB-4, fibrosis-4.

Between-group comparisons for secondary outcomes, including cardiovascular and non-cardiovascular death, were then performed. The incidence of cardiovascular death was as follows: Tertile 1, 4/67 (5%); Tertile 2, 2/67 (2%); and Tertile 3, 7/67 (10%; log-rank test, *P* = 0.17; Fig 1B, Table 2). In contrast, the incidence of non-cardiovascular death was as follows: Tertile 1, 3/67 (4%); Tertile 2, 3/67 (4%); Tertile 3, 19/67 (28%), which was significantly different between the groups (log-rank test, *P* < 0.001; Fig 1C, Table 2).

In subgroup analysis of the association between overall mortality and the FIB-4 index, consistent results were observed (Fig 2).

**Fig 2.**
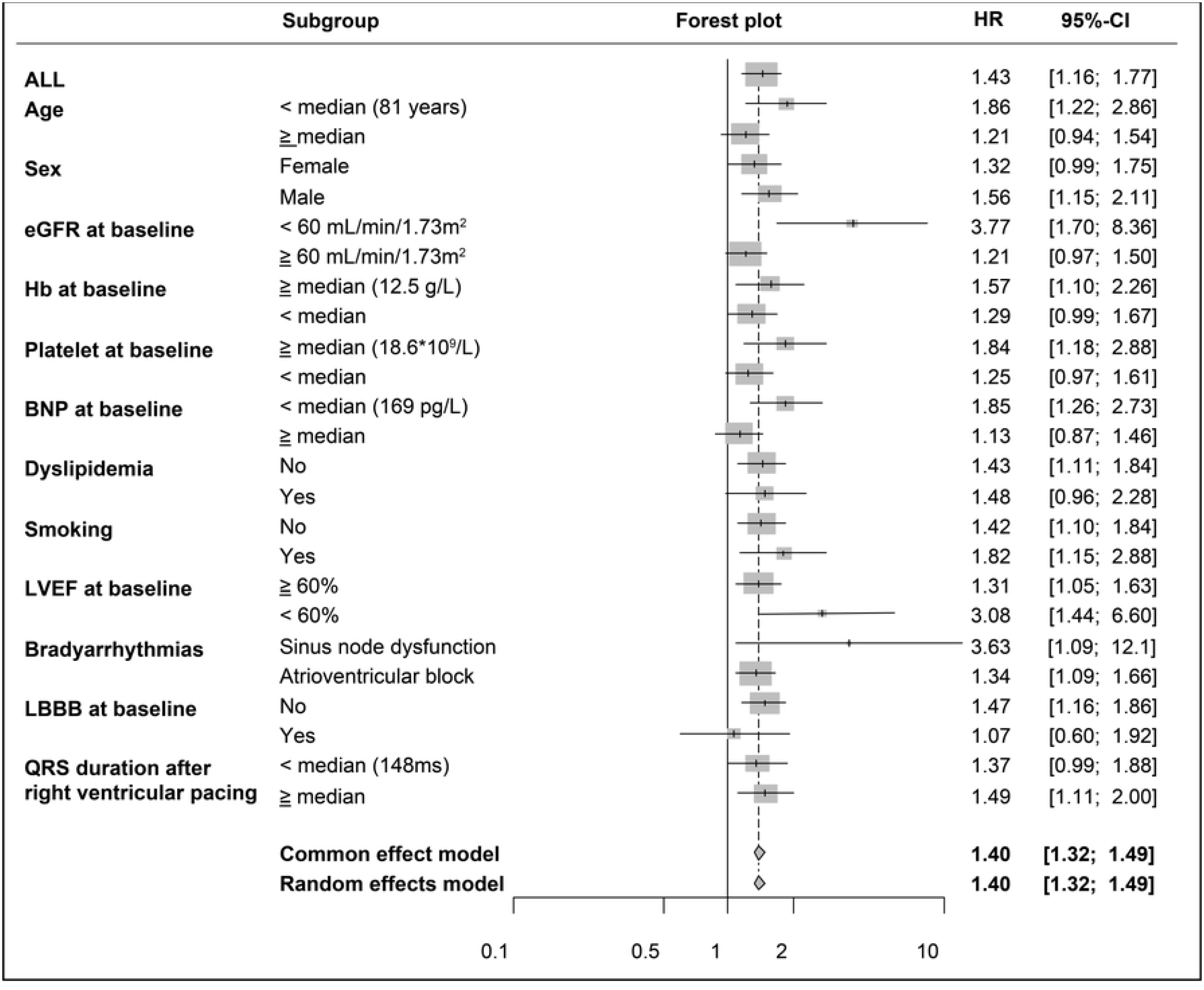
Forest plots from the subgroup analysis assessing the FIB-4 index and incidence of all-cause mortality. BNP, B-type natriuretic peptide; CI, confidence interval; eGFR, estimated glomerular filtration rate; FIB-4, fibrosis-4; Hb, hemoglobin; HR, hazard ratio; LBBB, left bundle branch block; LVEF, left ventricular ejection fraction.

In the Cox proportional hazards analysis for all-cause mortality, based on the FIB-4 index with Tertile 1 as the reference group, a significant increase in risk was observed in Tertile 3, with an unadjusted hazard ratio of 4.75 (95% CI: 2.05–11.0; *P* < 0.001; Table 3).

**Table 3.**
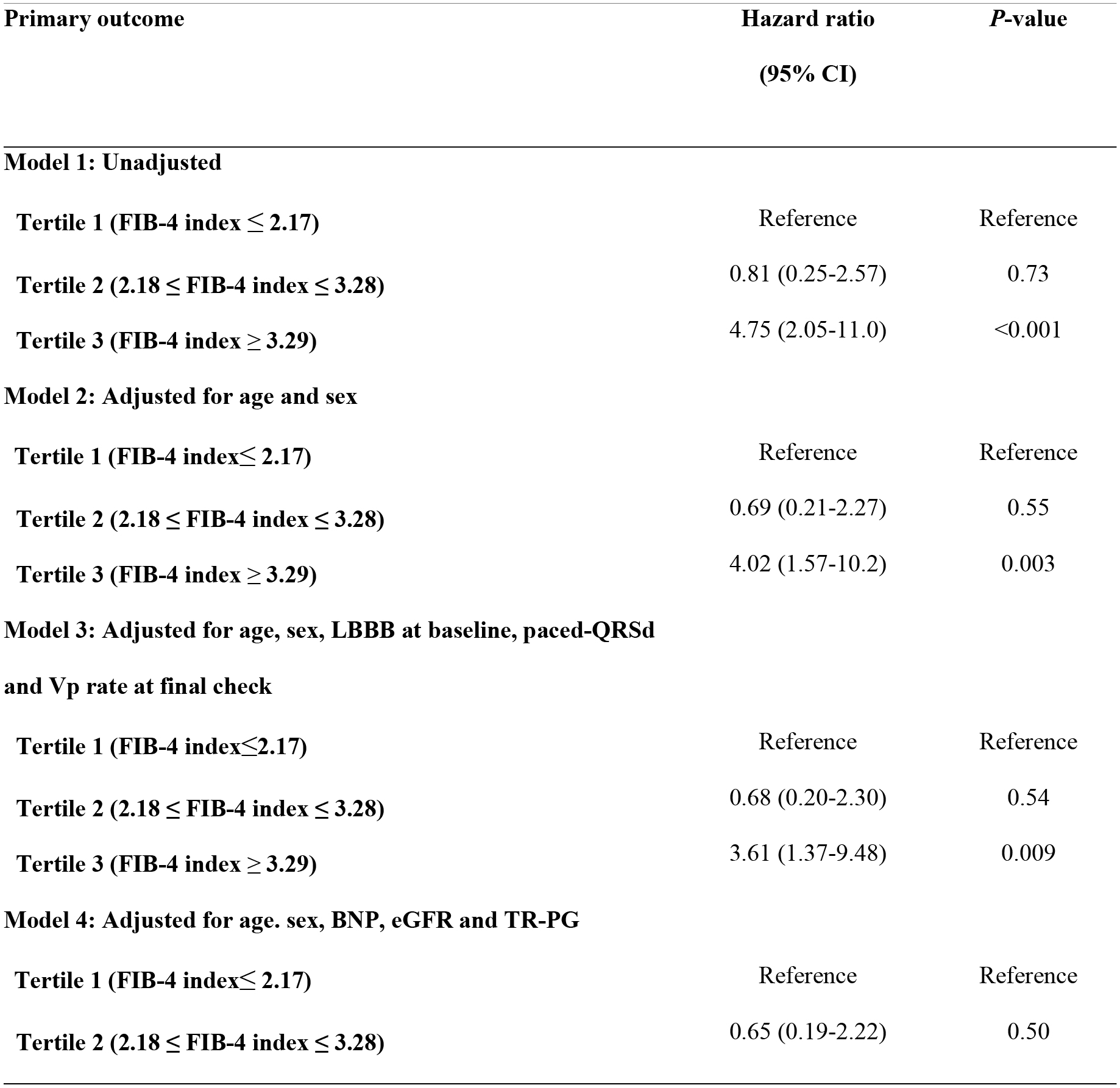
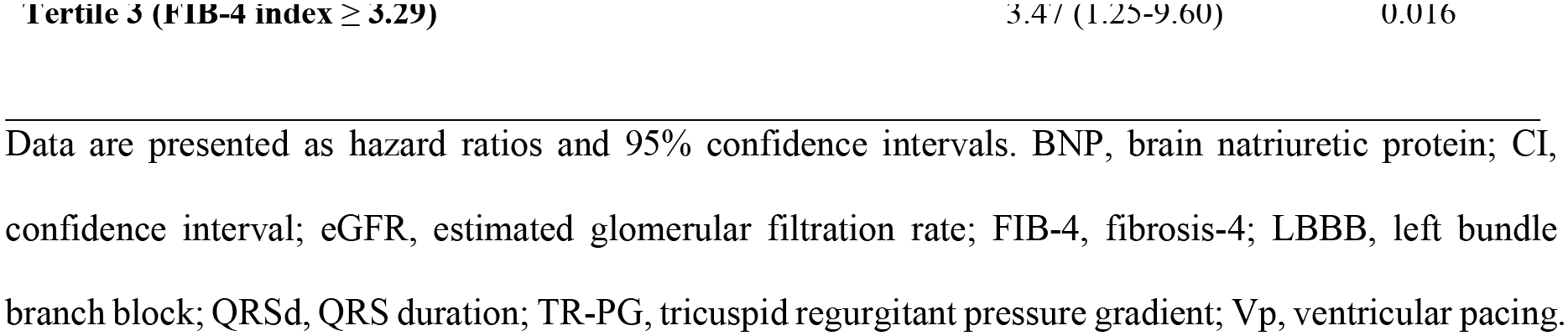
Cox proportional hazard analysis of the primary outcome.

Subsequently, the adjusted hazard ratio, considering clinically important factors, such as age, sex, and the presence or absence of LBBB at baseline; along with pacemaker-related factors, such as QRS duration during pacing and ventricular pacing rate, was 4.02 (95% CI: 1.57–10.2; *P* = 0.003) and 3.61 (95% CI: 1.37–9.48; *P* = 0.009) for Tertile 3 and Tertile 1, respectively (Table 3). In contrast, the adjusted hazard ratio considering factors related to congestion, such as BNP, eGFR, and TR-PG, was 3.47 (95% CI: 1.25–9.60; *P* = 0.016; Table 3).

Data are presented as hazard ratios and 95% confidence intervals. BNP, brain natriuretic protein; CI, confidence interval; eGFR, estimated glomerular filtration rate; FIB-4, fibrosis-4; LBBB, left bundle branch block; QRSd, QRS duration; TR-PG, tricuspid regurgitant pressure gradient; Vp, ventricular pacing.

The evaluation of the predictive accuracy of the FIB-4 index for all-cause mortality was performed using ROC curve analysis, resulting in an area under the curve of 0.72 (95% CI: 0.62– 0.82). The cut-off value for the FIB-4 index was 3.75, with a specificity of 0.86 and a sensitivity of 0.63 (Fig 3).

**Fig 3.**
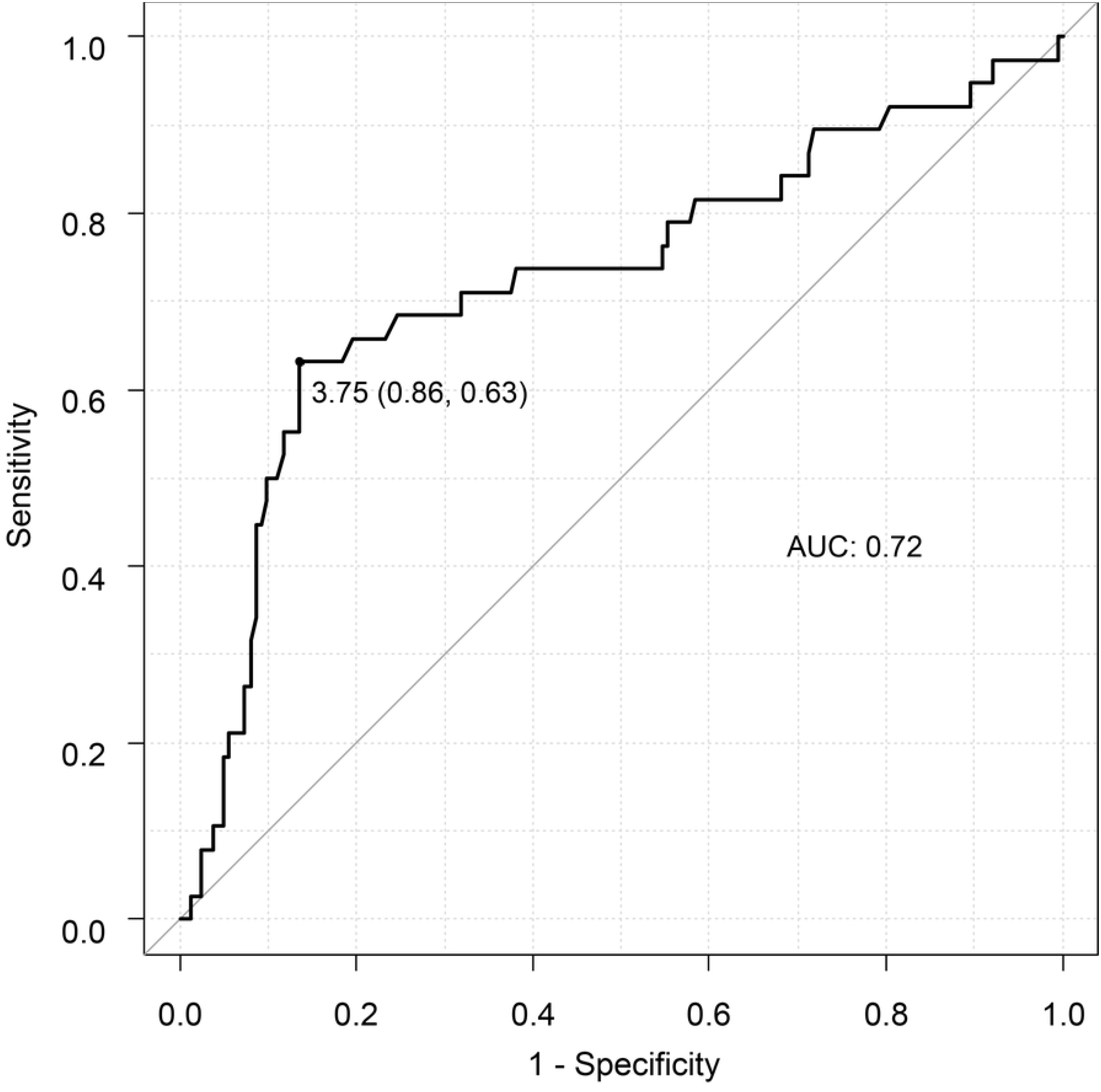
ROC curve analysis of the association between the FIB-4 index and the risk of all-cause mortality. ROC, receiver operating characteristic; AUC, area under the curve.

## Discussion

This study showed that in patients with RVP implants, a high FIB-4 index was associated with an increased risk of all-cause mortality. This trend remained consistent even after adjusting for pacemaker-related factors. Furthermore, a rightward nonlinear relationship between the FIB-4 index and all-cause mortality was demonstrated. These findings are this study’s novel contributions.

A meta-analysis compared cardiac resynchronization therapy and His/left bundle area pacing [13]; however, there is still a lack of studies specifically focusing on RVP and all-cause mortality. Spath et al. reported on a major composite endpoint, that included all-cause mortality, related to the lead position of RVP [14]; the all-cause mortality rate during the approximately 1000-day follow-up period was approximately 20% of the analyzed participants, which was similar to the death event rate in this study [14]. Subgroup analysis indicated that clinical outcomes improved when the QRS duration during pacing was narrow (< 130 ms) compared to when it was wide (≥ 130 ms) [14]. Therefore, we hypothesized that pacemaker-related factors may significantly impact mortality in patients with pacemakers and performed a multivariate analysis with pacing QRS duration and ventricular pacing rate as confounding factors. However, the results of our study revealed that the FIB-4 index was independently associated with all-cause mortality, regardless of these factors.

We also evaluated the association between the FIB-4 index and cardiovascular death as a secondary outcome. As mentioned earlier, RVP may significantly affect left ventricular contraction and valvular pathology due to its nonphysiological pacing pattern, increasing the risk of heart failure and atrial fibrillation [1–3]. Considering that the FIB-4 index may reflect the pathophysiology of circulatory failure, we hypothesized that there would be a significant association between the FIB-4 index and cardiovascular death. Although the Kaplan–Meier analysis showed no statistical significance, we believe that this might have been due to the extremely low number of cardiovascular death events during the 3-year follow-up period.

Notably, advancements in medical therapy for heart failure reduced ejection fraction, transcatheter aortic valve replacement [15], MitraClip procedures for valvular disease [16], and interventional treatments for myocardial infarction. These factors, along with the widespread adoption of ablation therapy for atrial fibrillation, indicate that long-term follow-up is necessary for further analysis of cardiovascular death. Therefore, although no statistical differences in cardiovascular death were observed, the results of future studies are anticipated to shed light on the clinical significance of the FIB-4 index.

This study has some limitations. It was a retrospective single-center study; therefore, the influence of potential biases, such as an imbalance in the patient population, cannot be denied. Moreover, owing to the limited number of deaths, a multivariate analysis with sufficient consideration of confounding factors could not be conducted. In addition, this study included patients with sinoatrial node dysfunction, in whom the degree of fusion pacing may be affected by the delay settings, which could potentially affect the clinical outcomes. Lastly, although the ventricular pacing rate is an important factor in clinical outcomes, this study only considered the pacing rate at the final follow-up. In reality, there may be long-term variations in the pacing rates for each patient, and it is necessary to investigate the relationship between temporal pacing rate changes and death events.

## Conclusion

In summary, the FIB-4 index may be a potential predictor of overall mortality in patients with right ventricular pacemakers.

## Data Availability

All relevant data are within the manuscript and its Supporting Information files.

## Acknowledgments

We are grateful to Dr. Takahiro Imaizumi for helpful discussions and comments on the manuscript.

## Authors’ Contributions

Naoya Inoue: Conceptualization, data curation, methodology, project administration, visualization, writing – original draft, writing – review & editing.

Shuji Morikawa: Formal analysis, writing – review & editing.

Takashi Ogane: Data curation, writing – review & editing.

Takehiro Hiramatsu: Data curation, conceptualization, formal analysis, investigation. writing – review & editing.

Toyoaki Murohara: Conceptualization, supervision, validation, writing – original draft, writing – review & editing.

## Supporting information

**S1 Table. Anonymous dataset of 201 patients**.

